# Age-stratified infection fatality rate of COVID-19 in the non-elderly informed from pre-vaccination national seroprevalence studies

**DOI:** 10.1101/2022.10.11.22280963

**Authors:** Angelo Maria Pezzullo, Cathrine Axfors, Despina G. Contopoulos-Ioannidis, Alexandre Apostolatos, John P.A. Ioannidis

## Abstract

The infection fatality rate (IFR) of COVID-19 among non-elderly people in the absence of vaccination or prior infection is important to estimate accurately, since 94% of the global population is younger than 70 years and 86% is younger than 60 years. In systematic searches in SeroTracker and PubMed (protocol: https://osf.io/xvupr), we identified 40 eligible national seroprevalence studies covering 38 countries with pre-vaccination seroprevalence data. For 29 countries (24 high-income, 5 others), publicly available age-stratified COVID-19 death data and age-stratified seroprevalence information were available and were included in the primary analysis. The IFRs had a median of 0.035% (interquartile range (IQR) 0.013 - 0.056%) for the 0-59 years old population, and 0.095% (IQR 0.036 - 0.125%,) for the 0-69 years old. The median IFR was 0.0003% at 0-19 years, 0.003% at 20-29 years, 0.011% at 30-39 years, 0.035% at 40-49 years, 0.129% at 50-59 years, and 0.501% at 60-69 years. Including data from another 9 countries with imputed age distribution of COVID-19 deaths yielded median IFR of 0.025-0.032% for 0-59 years and 0.063-0.082% for 0-69 years. Meta-regression analyses also suggested global IFR of 0.03% and 0.07%, respectively in these age groups. The current analysis suggests a much lower pre-vaccination IFR in non-elderly populations than previously suggested. Large differences did exist between countries and may reflect differences in comorbidities and other factors. These estimates provide a baseline from which to fathom further IFR declines with the widespread use of vaccination, prior infections, and evolution of new variants.

**Highlights:** *Across 31 systematically identified national seroprevalence studies in the pre-vaccination era, the median infection fatality rate of COVID-19 was estimated to be 0.035% for people aged 0-59 years people and 0.095% for those aged 0-69 years.

*The median IFR was 0.0003% at 0-19 years, 0.003% at 20-29 years, 0.011% at 30-39 years, 0.035% at 40-49 years, 0.129% at 50-59 years, and 0.501% at 60-69 years.

*At a global level, pre-vaccination IFR may have been as low as 0.03% and 0.07% for 0-59 and 0-69 year old people, respectively.

*These IFR estimates in non-elderly populations are lower than previous calculations had suggested.

## 1. INTRODUCTION

The Coronavirus Disease 2019 (COVID-19) pandemic has had grave worldwide consequences. Among people dying from COVID-19, the largest burden is carried by the elderly (1), and persons living in nursing homes are particularly vulnerable (2). However, non-elderly people represent the vast majority of the global population, with 94% of the global population being younger than 70 years old, 91% being younger than 65 years old, and 86% being younger than 60 years old. It is therefore important to get accurate estimates of the infection fatality rate (IFR) of COVID-19 among non-elderly people, i.e., the proportion of deceased among those infected, and to assess the age-stratification of IFR among non-elderly strata. Such assessments carry profound implications in public health, from evaluating the pertinence of prevention measures to vaccine strategies. Several previous evaluations (3-6) have already synthesized information on age-stratified estimates of IFR. Most of those used data from early published studies, and these tended to have information from mostly hard hit countries, thus potentially with inflated IFR estimates. Moreover, several analytical and design choices for these reviews and data syntheses can be contested (7) and many more potentially informative seroprevalence studies have been published since then. We recently examined age stratified IFR in the non-elderly populations as a secondary analysis of a project focused primarily on the IFR in the elderly (8); however, in this evaluation only studies with sampling until the end of 2020 and which had a large number of elderly individuals were considered. The median IFR considering available data from fully representative general population studies was 0.0009% at 0-19 years, 0.012% at 20-29 years, 0.035% at 30-39 years, 0.109% at 40-49 years, 0.34% at 50-59 years, and 1.07% at 60-69 years without accounting for seroreversion (loss of antibodies over time in previously infected individuals)Including also convenience sample studies (and again without accounting for seroreversion) the respective age groups median IFR estimates were 0.001%, 0.010%, 0.023%, 0.050%, 0.15%, and 0.49%.(8).

Here, we extended the analysis of COVID-19 IFR in non-elderly age-strata pertaining to the pre-vaccination era to examine studies published until mid-2022 regardless of whether they had many elderly participants as well, while using rigorous methods for study selection and analysis. We focused on studies that evaluated seroprevalence in representative general population samples at a national level. We also explored whether population and other features were associated with the IFR in the non-elderly population.

## 2. METHODS

### 2.1 Design and protocol

This was a mixed-methods analysis combining data from different sources. Analyses of IFR estimation in the non-elderly were performed in countries where information on age-stratified COVID-19 deaths was available so as to be able to separate deaths among the non-elderly. The protocol for this study was registered at the Open Science Framework (https://osf.io/xvupr) prior to full data analysis but after piloting data availability and after having done analyses on some studies as part of a related project focused on IFR estimates in the elderly (8). A secondary project using similar search strategies and eligibility criteria but focusing on relative seroprevalence ratios in different age groups (9) has been included in the same protocol and published separately.

### 2.2 Eligible seroprevalence studies

We identified seroprevalence studies (peer-reviewed publications, official reports, or preprints) in the live systematic review SeroTracker (10) and performed a PubMed search using the string “seroprevalence AND (national OR stratified) AND COVID-19” to identify potentially eligible studies that were recently published and thus may not have been yet indexed in SeroTracker. The initial search was performed on February 8, 2022 and updated on May 25, 2022.

We included only those studies on SARS-CoV-2 seroprevalence that met the following criteria:

i. Sampled any number of participants aged <70 years in a national representative sample.
ii. Sampling was completed by end February, 2021 and at least 90% of the samples had been collected before the end January 2021 (to avoid the impact of vaccination on IFR calculations).
iii. Adults (≥21 years old) were included, regardless of whether children and/or adolescents were included or not.
iv. Provided an estimate of seroprevalence for non-elderly people (preferably for <70 years and/or <60 years, but any cut-off between 54 and 70 years was acceptable)
v. Explicitly aimed to generate samples reflecting the general population.

We excluded studies focusing on patient cohorts (including residual clinical samples), blood donors, workers (healthcare or other), and insurance applicants and studies where the examined population might have had lower or higher risk than the general population, as explained and justified elsewhere (9).

Similar to the respective protocol for estimating IFR in the elderly (8) and the project on seroprevalence ratios in non-elderly vs elderly (9), we used predefined rules (i) for studies done in the USA (only those that had adjusted the seroprevalence estimates for race/ethnicity were retained, since this factor is known to associate strongly with the risk of SARS-CoV-2 infection); (ii) for studies with several sampled (sub)regions of a country (we accepted those where the sampling locations were dispersed across the country to form a reasonable representation of the entire country); (iii) for studies where crude seroprevalence was less than [1-test specificity] and/or the 95% confidence interval of the seroprevalence went to 0% (excluded, since the uncertainty on seroprevalence [and thus also IFR] for them was very large); and (iv) for age boundaries (excluded studies that included in their sampling only children and/or adolescents without any adults 21 years or older; otherwise studies were accepted regardless of presence or not of upper or lower boundaries).

Finally, the main analyses considered only studies from countries where information was available on the proportion of cumulative COVID-19 deaths among non-elderly with an upper cutoff placed between 60-70 years. Countries without this information were considered in sensitivity analyses while making certain assumptions for imputation of the age distribution of COVID-19 deaths (as discussed below).

### 2.3 Extracted information

Data extraction for eligible articles was performed in duplicate by at least two authors independently (AA, AMP, DCI) and disagreements were discussed. In cases of persistent disagreements, a third author arbitrated.

For each potentially eligible study, we tried to identify available data on the proportion of cumulative COVID-19 deaths among people <70 years old and among people <60 years old, which are the two main definitions for the non-elderly population in our analysis. If data were not available for these two cut-offs, but were available for a cut-off of <65, we imputed the respective death data for cut-offs of <70 and <60. For the imputations, we assumed that in a 10-year interval in that age vicinity, 1/3 of the deaths had occurred in the lower 5-year bin and 2/3 of the deaths had occurred in the upper 5-year bin. For example, if data on deaths were given for the age bins <55, 55-65 and 65-75 years, we assumed that 1/3 of the deaths in the age bin 55-65 occurred in the 55-60 years group so as to estimate deaths <60 years; and we assumed that 2/3 of deaths in the age bin 65-75 occurred in the 65-70 years group so as to estimate deaths <70 years. Studies done in countries where there was no available information on age-stratified COVID-19 deaths with an age-cutoff in the 60-70 range were considered only in sensitivity analyses with imputation of age distribution of COVID-19 deaths (as discussed below).

Similar to previous projects (3, 9), we extracted from all eligible seroprevalence studies their information on country, recruitment and sampling strategy, dates of sample collection, sample size in the non-elderly group (using age cutoffs <70, <65, and <60, whichever were available), and types of SARS-CoV2 antibodies measured (immunoglobulin G (IgG), IgM and IgA).

For the non-elderly population, we extracted the estimated unadjusted seroprevalence (positive samples divided by all samples tested), the most fully adjusted seroprevalence, and the factors that the authors considered for adjustment in the most fully adjusted calculations.

Antibody titers may decline over time. For example, a modelling study estimating the average time from seroconversion to seroreversion at 3-4 months (11) and other investigators have also found steep decreases in antibody assay sensitivity over time (12) and a systematic review found large variability in seroreversion rates across assays and studies (13). Therefore, for consistency, if there were multiple different time points when seroprevalence was assessed in a given study, we selected the one that gave the highest seroprevalence estimate and when there was a tie we chose the earliest one (in a sensitivity analysis, we excluded from the calculations studies where the chosen time point was not the latest).

Whenever authors had already adjusted for seroreversion, we used the seroreversion-adjusted estimate. When the authors had not adjusted for seroreversion, we adjusted for 5% monthly rate of seroreversion, correcting the observed seroprevalence by 0.95^m^-fold, where m is the number of months from the peak of the first epidemic wave in the specific location. The peak of the first epidemic wave was defined as one week before the date with the highest rolling average 7-day mortality (according to Worldometer) until August 31, 2020. If two or more dates were tied for peak values, we chose the date corresponding to the midpoint between the first and last one.

Whenever authors had not adjusted for antibody test performance (sensitivity and specificity), we used the Gladen-Rogan formula (14) to make this adjustment.

The population size overall and in the non-elderly population (using cut-offs of 70 years and of 60 years) in the relevant country were primarily obtained from the seroprevalence study. If not provided in the study, we used either populationpyramid.net, official population data (e.g., the latest available national census), or worldpopulationreview.com, in that order, to retrieve the relevant number for the end of 2020 (or as close as possible to that date).

Cumulative COVID-19 deaths overall and in the non-elderly population (using separately the <70 and <60 year cut-offs) for the relevant country were extracted, whenever available, from COVerAGE-DB (15) [https://osf.io/mpwjq/], The Demography of COVID-19 Deaths database of Institut national d’études démographiques (DCD-INED) (16) [https://dc-covid.site.ined.fr/en/], official reports, or Worldometer, in that order. Both COVerAGE-DB and DCD-INED are compilations of official reports. The total number of deaths (confirmed and probable) was preferred whenever available. We extracted the accumulated deaths until the date 1 week after the midpoint of the seroprevalence study period (or the date closest to this that had available data) to account for different delays in developing antibodies versus dying from infection. For a sensitivity analysis, we extracted data on accumulated deaths until the date 2 weeks after the midpoint. By midpoint, one refers to the median date of sampling, or (if the rate of sampling over time is unclear and there is no suggestion that it was uneven in different time periods) the time point that is equidistant from the start and end dates. If the seroprevalence study claimed strong arguments to use another time point or approach, while reporting official statistics on the number of COVID-19 deaths overall and in the non-elderly population, we extracted that number instead. The number of deaths is only an approximation and may be biased for various reasons, including different time lag from infection to death and imperfect diagnostic documentation of COVID-19 potentially leading to either under-or over-counting (17).

### 2.4 Estimation of the number of infected and deceased non-elderly

The number of infected people was estimated by multiplying the adjusted estimate of seroprevalence and the population size in non-elderly. If a study did not give an adjusted seroprevalence estimate, we used the unadjusted seroprevalence instead, as mentioned above. Both adjusted and unadjusted estimates were corrected for test performance and seroreversion, unless already corrected by the authors. For locations that did not report seroprevalence data for the non-elderly group for the <60 and <70 cut-offs, we used the seroprevalence estimate for the closest cut-off available in the 60-70 range. We applied a correction for studies that excluded persons with diagnosed COVID-19 from participating in their sample, primarily using study authors’ corrections (e.g., PCR tests) or adding the number of identified COVID-19 cases in community-dwelling non-elderly for the location until the seroprevalence study midpoint. For studies that performed surveys using both seroprevalence and PCR testing and presented as main analyses data for being positive in either test, we used the data that reflect infection documented with either way.

The total number of COVID-19 fatalities in non-elderly (for the <60 and <70 cut-offs) were counted from available sources until 1 week after the midpoint of the seroprevalence study period. If the age distribution of COVID-19 deaths was only available for a date more than 1 week apart from the preferred one, we assumed that the proportions of age-stratified deaths were stable between the time points and inferred the total number of fatalities for the preferred date. That is, we calculated the percentage of fatalities in non-elderly for the available date (namely, the number of deaths in non-elderly divided by total number of deaths) and multiplied it with the total number of deaths for the preferred date to obtain the COVID-19 fatalities in non-elderly for the preferred date. When COVID-19 deaths were not available for the <60 and <70 cut-offs (e.g. given only for the age bin 65-75), we imputed them using the 1/3-rule imputation for breaking down 10-year bins to 5-year bins, as mentioned above.

### 2.5 IFR estimation

We calculated the inferred IFR in the non-elderly, by dividing the number of deaths in this population group by the number of infected people for the same population group. We performed separate calculations defining the non-elderly as those being <60 and those being <70 years old.

### 2.6 Data extraction for age-stratified analyses within the non-elderly group

The same considerations outlined above for the entire non-elderly population were applied for extracting information on seroprevalence, population size and the number of COVID-19 deaths for separate age strata bins within the non-elderly population, whenever available. Whenever seroprevalence estimates and COVID-19 mortality data were available for specific granular age groups, we complemented data extraction for all available age strata.

Studies were excluded from the age-stratified analysis if no mortality data were available for any age stratum of maximum width 20 years and maximum age 70 years. We used the same time points as those selected for the overall non-elderly data analysis. We included all age strata with a maximum width of 20 years and available COVID-19 mortality information.

We corresponded the respective seroprevalence estimates for each age stratum with eligible mortality data. Consecutive strata of 1-5 years were merged to generate 10-year bins. For seroprevalence estimates we used the age strata that most fully cover/correspond to the age bin for which mortality data are available; specifically for the youngest age groups, seroprevalence data from the closest available group with any sampled persons ≤20 years were accepted. E.g. for the Ward et al UK study (18), the youngest stratum with seroprevalence data is 18-24 years old. Population statistics for each analyzed age bin were obtained from the same sources as for the overall analysis for the non-elderly.

For countries for which age information was missing for a proportion of the cumulative COVID-19 deaths, we assumed the age distribution to be the same as for the non-missing proportion.

### 2.7 Data synthesis

The main outcomes were the IFR in people <60 years old and <70 years old, as well as age-stratified IFR estimates in smaller age bins among the non-elderly.

Similar to previous work on IFR-estimating studies (3,8), we estimated the sample size-weighted IFR of non-elderly (separately for <60 and <70 years old) for each country (if multiple studies were available for that country) and then estimated the median and range of IFRs across countries. We expected very large heterogeneity among IFR estimates, therefore we did not use meta-analysis methods.

To generate plots of IFRs with some estimates of uncertainty, we performed calculation of 95% CIs of IFRs based on extracted 95% CIs from seroprevalence estimates. Primarily, 95% confidence intervals are direct extractions from the seroprevalence studies. For studies that did not report such intervals, we complemented the analysis with a calculation using the number of sampled and seropositive non-elderly individuals (Clopper Pearson interval calculation). For those that provided adjusted estimates for age brackets, we combined estimates for each study using a fixed effects inverse variance meta-analysis (of arcsine transformed proportions) to obtain 95% CIs. No further factors were introduced in the calculation beyond the adjustments made by seroprevalence study authors (except adjusting estimates for test performance using the Gladen-Rogan formula and adjusting also for seroreversion -assuming 5% monthly seroreversion-, where applicable).

Similar to the overall non-elderly analyses, for age strata with multiple estimates from the same country, we calculated the sample size-weighted IFR per country before estimating median IFRs across countries for age groups 0-19, 20-29, 30-39, 40-49, 50-59, and 60-69 years. IFR estimates were placed in these age groups according to their midpoint, regardless of whether they perfectly match the age group or not, e.g. an IFR estimate for age 18-29 years was placed in the 20-29 years group. As for the main analysis, whenever no adjustment had been made for test performance, we adjusted the estimates for test performance using the Gladen-Rogan formula; and whenever there had been no adjustment for seroreversion, we corrected the results assuming 5% monthly seroreversion.

### 2.8 Sensitivity analyses

We performed the following sensitivity analyses

1. Limited to high-income countries. Under- and over-counting of deaths may occur also in high-income countries (17), but the concern for under-counting is more serious in other countries (19). Nevertheless, under-counting may be much less of a problem in non-elderly people than in the elderly.
2. Considering deaths up to 2 weeks after the midpoint of seroprevalence sampling, instead of just one week.
3. Excluding studies where the chosen time point was not the latest available (observed seroprevalence has declined subsequently).
4. Exploring different seroreversion corrections of the IFR by X^m^-fold, where m is the number of months from the peak of the first epidemic wave in the specific location.X was given values of 1.00, 0.99, and 0.90 corresponding to no seroreversion, 1%, and 10% relative rate of seroreversion every month from the peak of the first epidemic wave in the specific location to the date of seroprevalence estimate.
5. Including in the overall calculations of IFR in the non-elderly also imputed data from countries where the proportion of COVID-19 deaths occurring among the non-elderly was not available. This is a post-hoc sensitivity analysis and it was adopted because a substantial number of studies fell in this category. Specifically, we assumed that the proportion of COVID-19 deaths represented by the non-elderly was a minimum of 10% for 0-59 years (and 20% for 0-69 years) and a maximum of 60% for 0-59 years (and 90% for 0-69 years).

### 2.9 Evaluation of heterogeneity

We explored whether the estimated IFR for the non-elderly across different countries was associated with the structure of the age pyramid in the population of each country. Specifically, we performed meta-regression analyses of the country IFR in the non-elderly against the proportion of the non-elderly population that is <50 years old. Separate regression analyses were performed using the definition of non-elderly as being <70 years old and <60 years old.

Additional factors that were explored for association with IFR in the non-elderly were country-income (high-income country versus other), and the population-level annual mortality rate in each country (https://worldpopulationreview.com/country-rankings/death-rate-by-country). We used these observations in trying to extrapolate to the respective features of the global population, to try to approximate the IFR among the non-elderly in the global population.

## 3. RESULTS

### 3.1 Eligible studies

By February 8, 2022, Serotracker had 2930 seroprevalence studies, of which 547 entries were described as “national”. Of those, 420 had their sampling end date before February 28, 2021. 183 were characterized as “household and community samples” or “multiple populations”. Of those, 107 were of low, moderate or unclear risk of bias. We screened in-depth the 107 entries and 73 were excluded. Therefore, 34 studies were eligible from this source. Our search on PubMed yielded 474 items, of which four additional eligible studies were identified. On May 25, 2022, we updated the search and found 2 additional studies to be included. In total, data from 40 studies which covered national seroprevalence estimates for 38 different countries were extracted and analyzed (18,20-58). 30 countries had publicly available age-stratified COVID-19 death data. The report of one of these countries (Austria) did not report any information on age-stratified seroprevalence. Therefore, 29 countries with data from 31 studies were included in the primary analysis (Appendix Figure 1).

### 3.2 Characteristics of eligible studies

Table 1 shows the main characteristics for the 31 studies with publicly available age-stratified COVID-19 death and seroprevalence data. As shown, these data originated from 24 high income countries and 5 other countries.

**Table 1.**
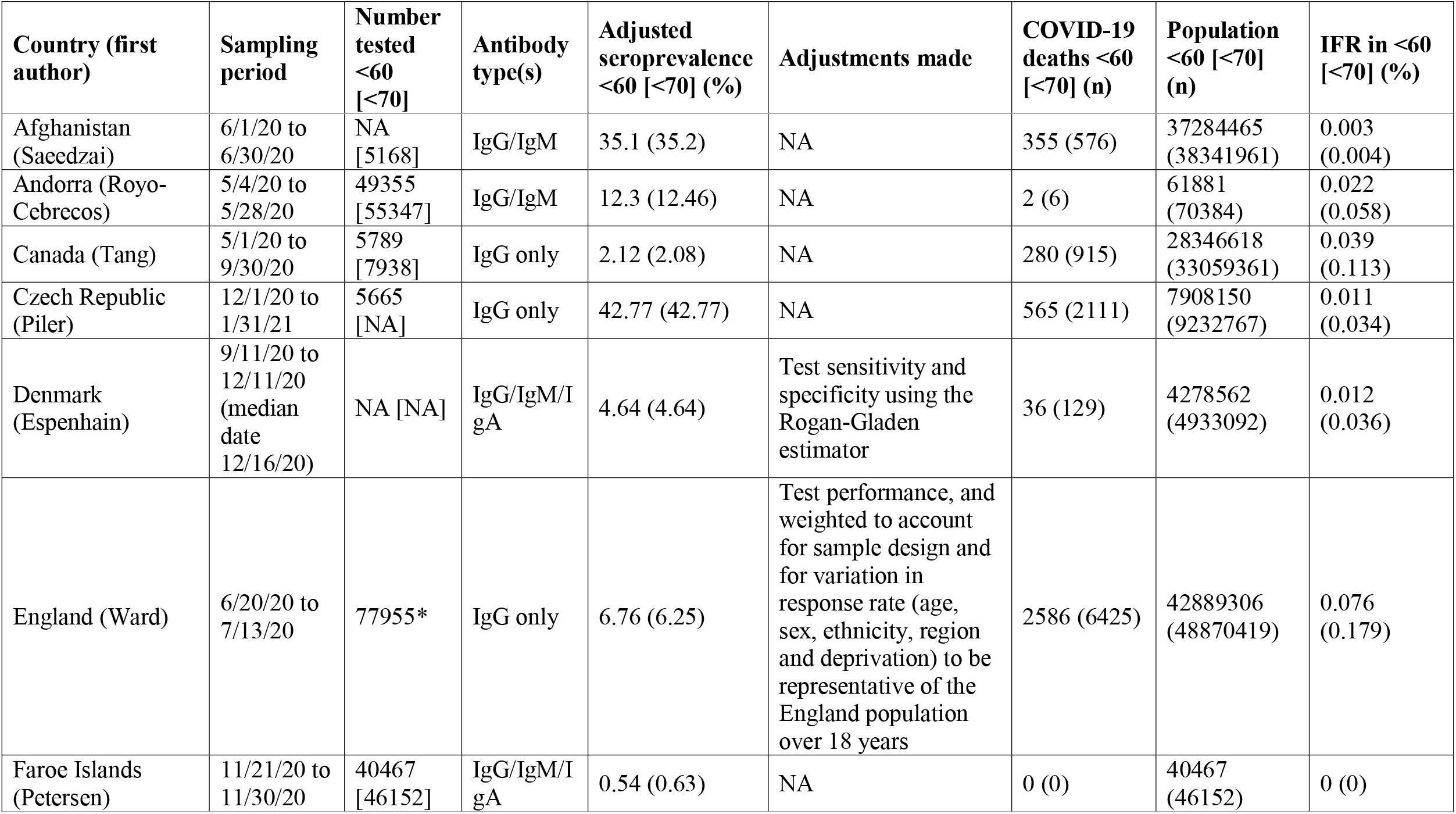
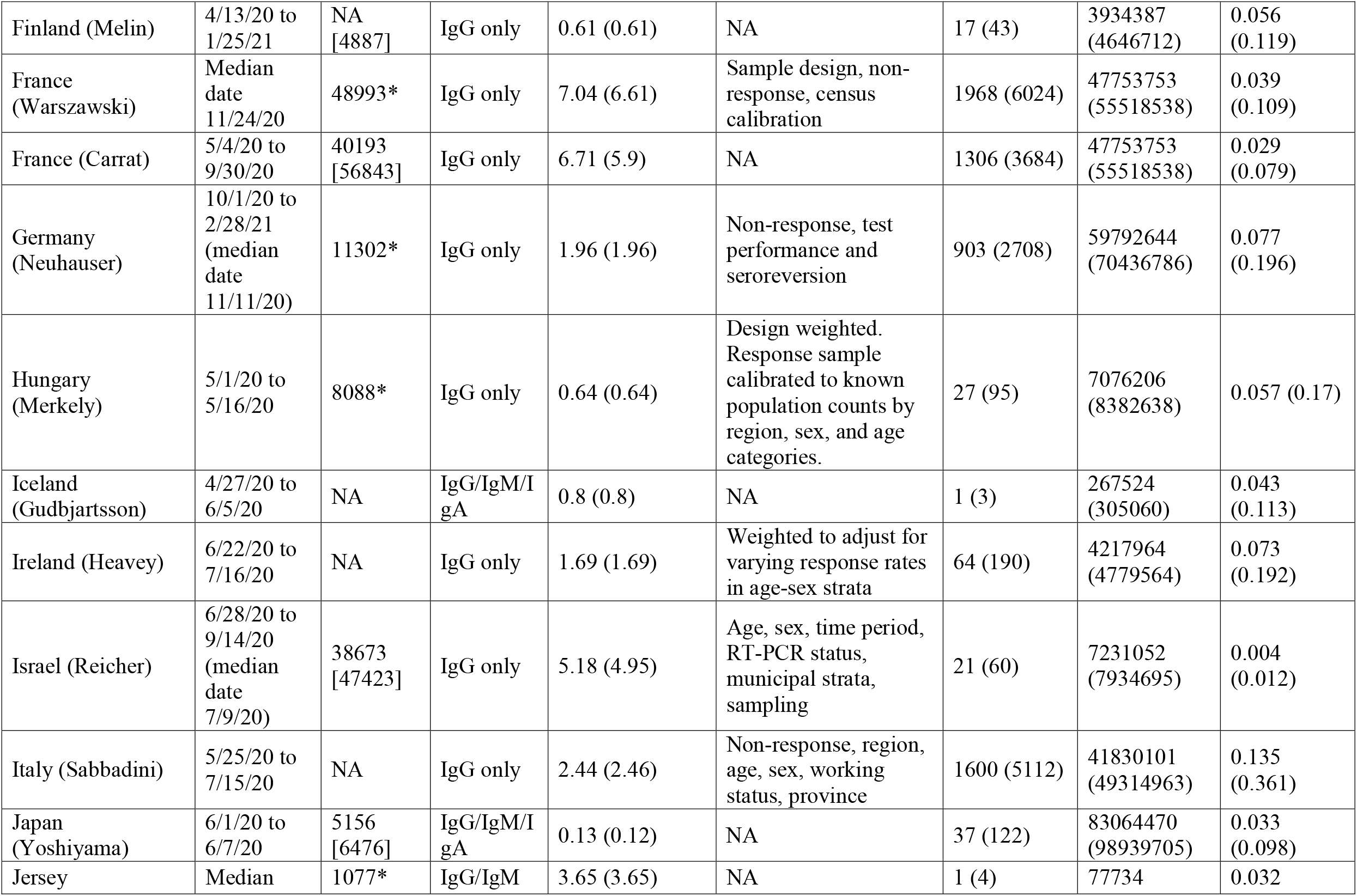
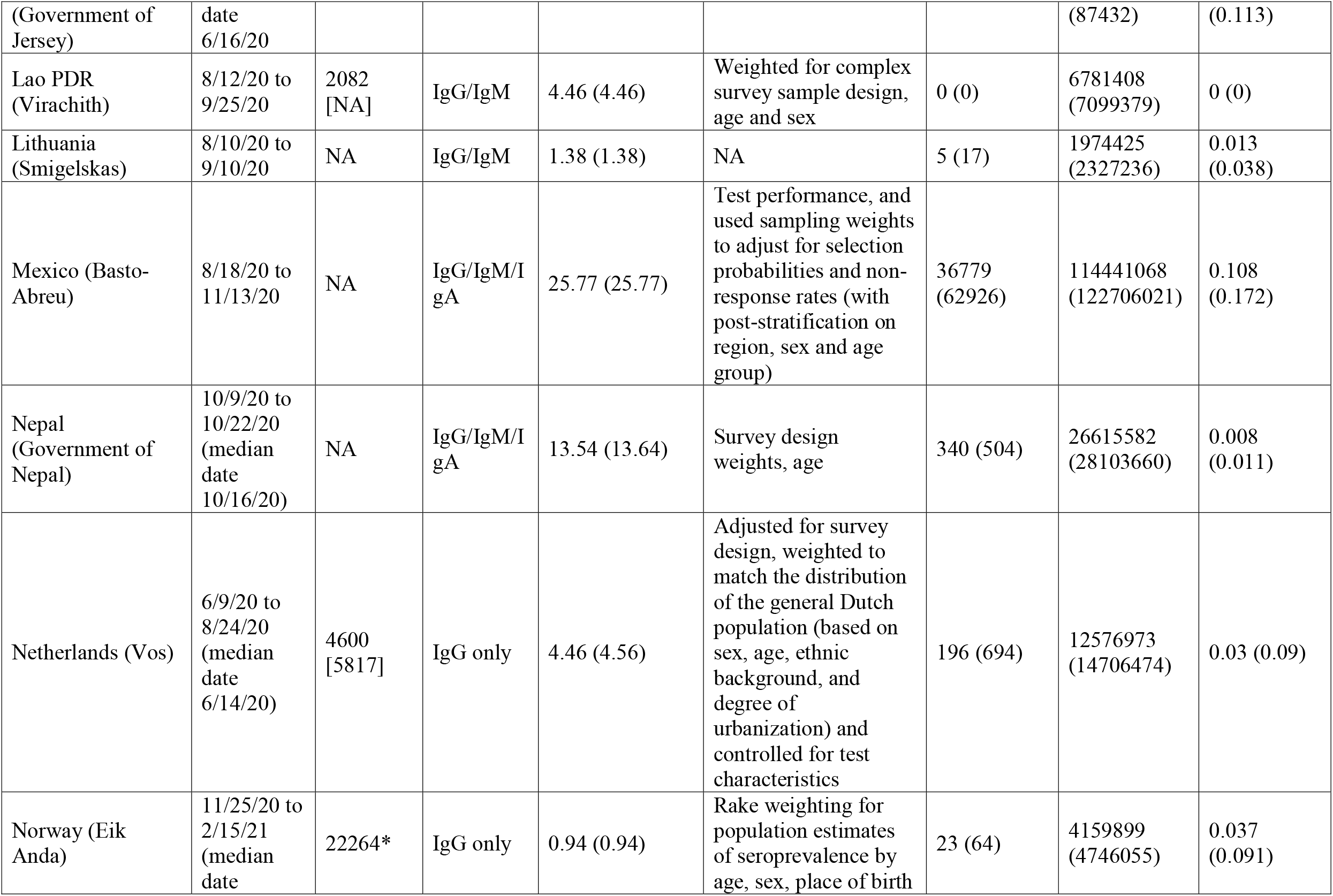
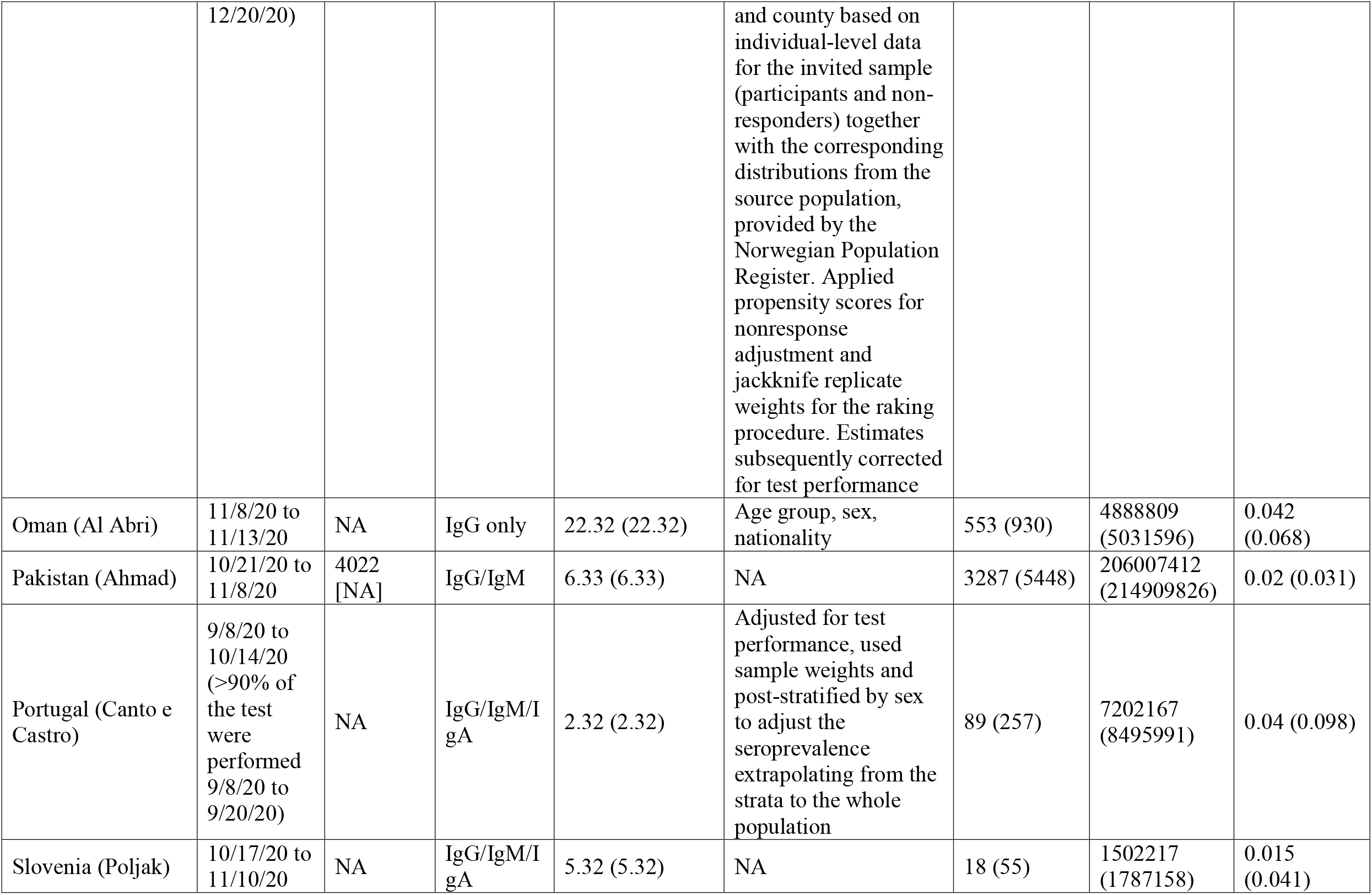
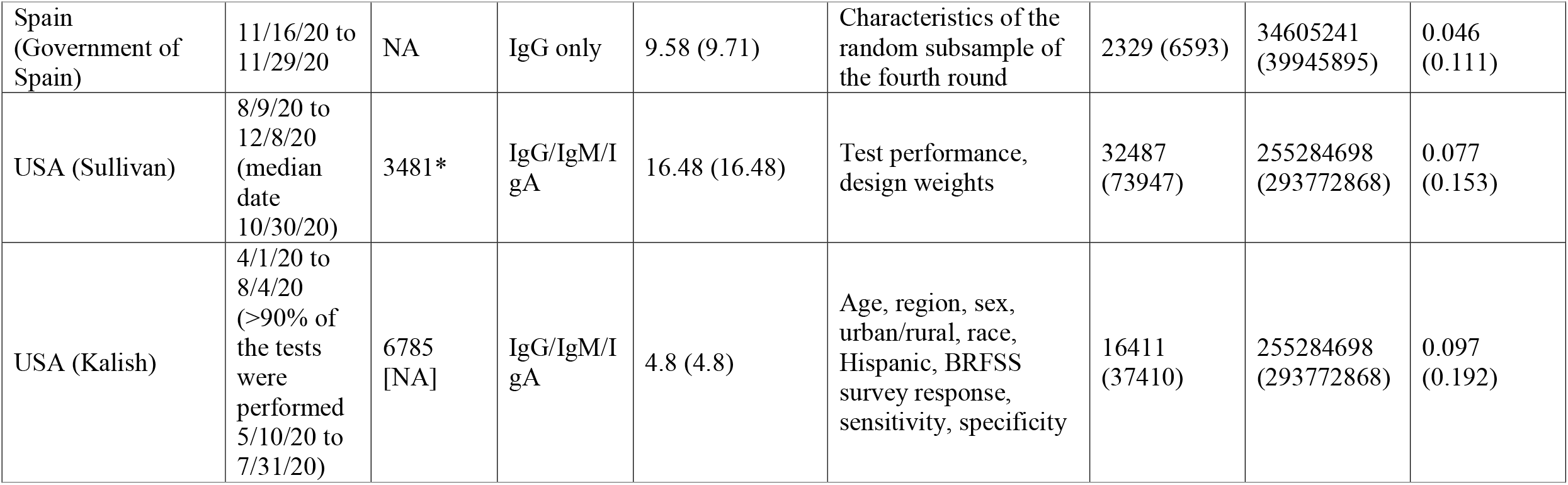
Eligible studies for the main analysis (those countries that have age-stratified COVID-19 death data and seroprevalence information)

### 3.3 IFR estimates in the non-elderly

In 29 countries of the primary analysis, with age-stratified COVID-19 death and seroprevalence data, IFRs in non-elderly (Figure 1, Table 1) had a median of 0.035% (interquartile range (IQR) 0.013 - 0.056%, Figure 1A) for the 0-59 years old population, and of 0.095% (IQR 0.036 - 0.125%, Figure 1B) for the 0-69 years old population. Figure 1 also shows 95% CIs for IFRs based on 95% CIs for seroprevalence estimates.

**Figure 1.**
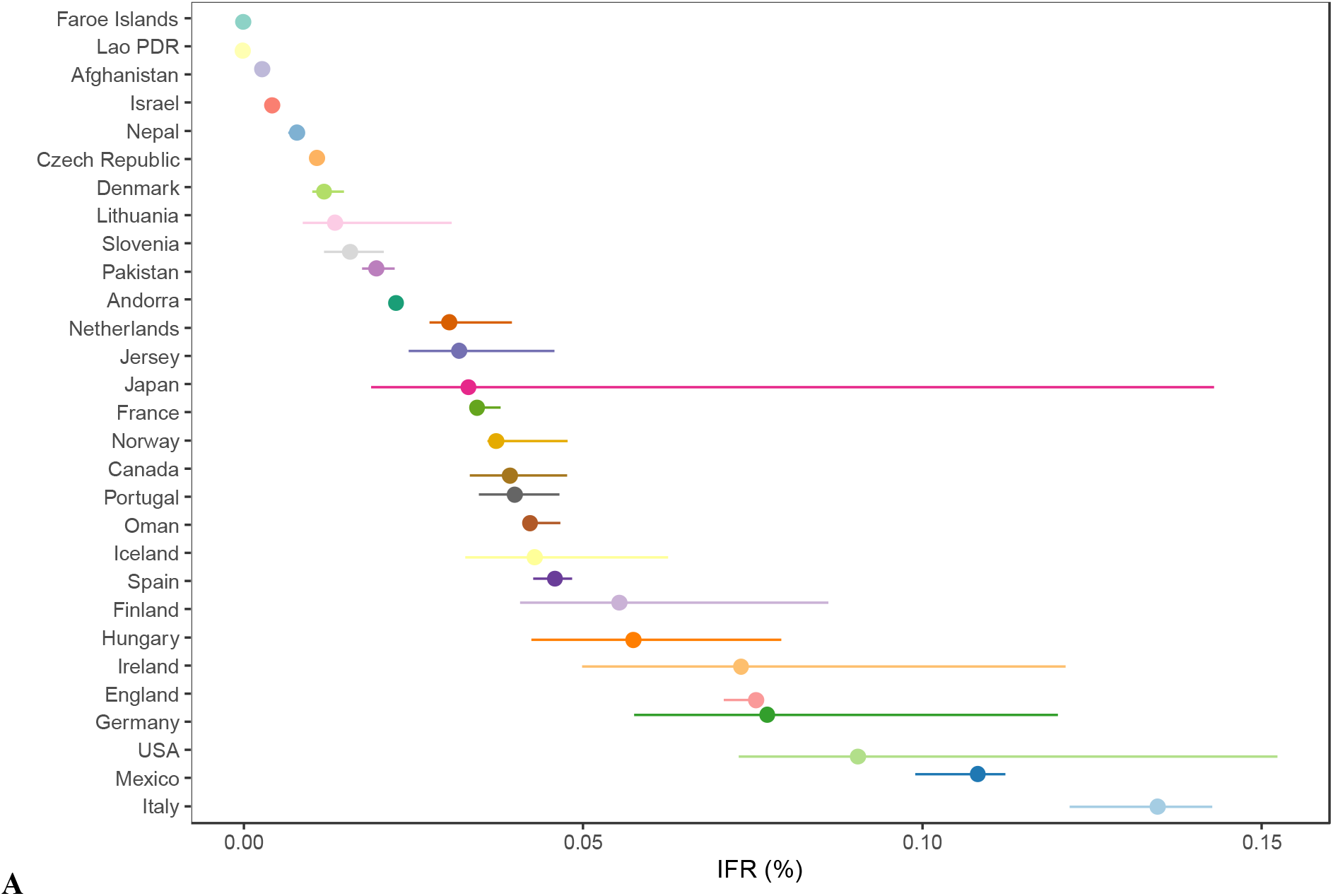
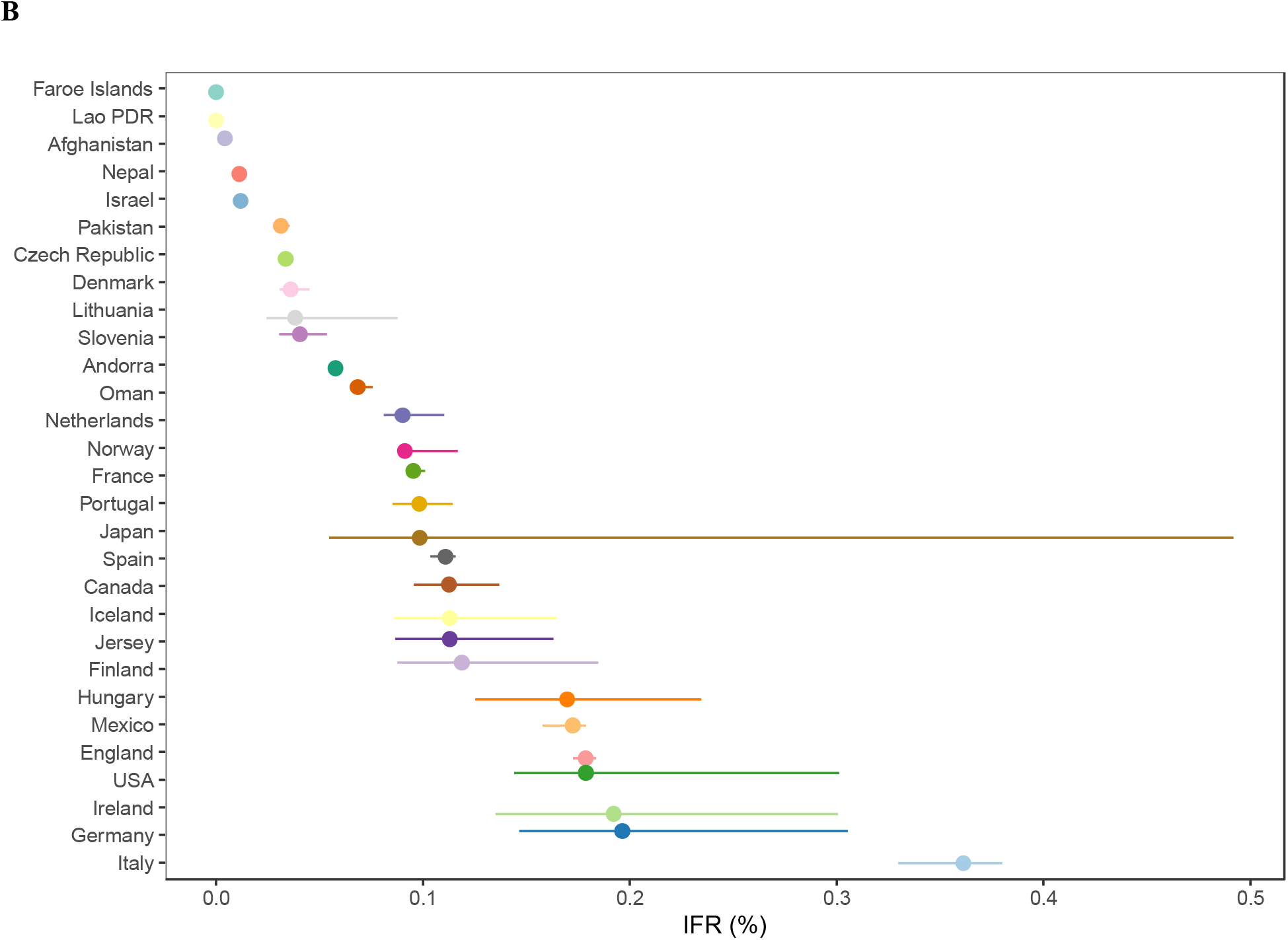
Infection fatality rate (IFR) and 95% confidence interval per country. Panel (A) for <60 years old people. Panel (B) for <70 years old people. Note: For multiple estimates from the same country (France and USA), we calculated the sample size-weighted IFR per country. The 95% confidence intervals are estimated primarily as direct extractions from the seroprevalence studies. For studies that did not report 95% confidence intervals, we complemented with a calculation using the number of sampled and seropositive individuals. For those that provided adjusted estimates for age brackets (e.g., 0–9, 10–19, 20-29, etc.), we combined estimates for each study using a fixed effects inverse variance meta-analysis (of arcsine transformed proportions) to obtain 95% confidence intervals. Asymmetry around point estimates may be observed for these cases, since point estimates were calculated by multiplying age bracket seroprevalence by the corresponding population count (which is preferable, since it takes into account population distribution). Please note that uncertainty in seroprevalence estimates is larger than conveyed by typical 95% confidence intervals.

### 3.4 IFR estimates per narrow age strata

For the narrow age bins analysis (Figure 2), the median IFR was 0.0003% (IQR, 0.0000 to 0.002) at 0-19 years, 0.003% (IQR, 0.000 to 0.007) at 20-29 years, 0.011% (IQR, 0.005 to 0.031) at 30-39 years, 0.035% (IQR, 0.011 to 0.077) at 40-49 years, 0.129% (IQR 0.047 to 0.220) at 50-59 years, and 0.501% (IQR, 0.208 to 0.879) at 60-69 years. Excluding from the calculations age bins with 0 deaths (where IFR is thus calculated as 0.000% but has very large uncertainty), the median IFR was 0.001%, 0.006%, 0.012%, 0.048%, 0.158%, and 0.544% in these age bins, respectively.

**Figure 2.**
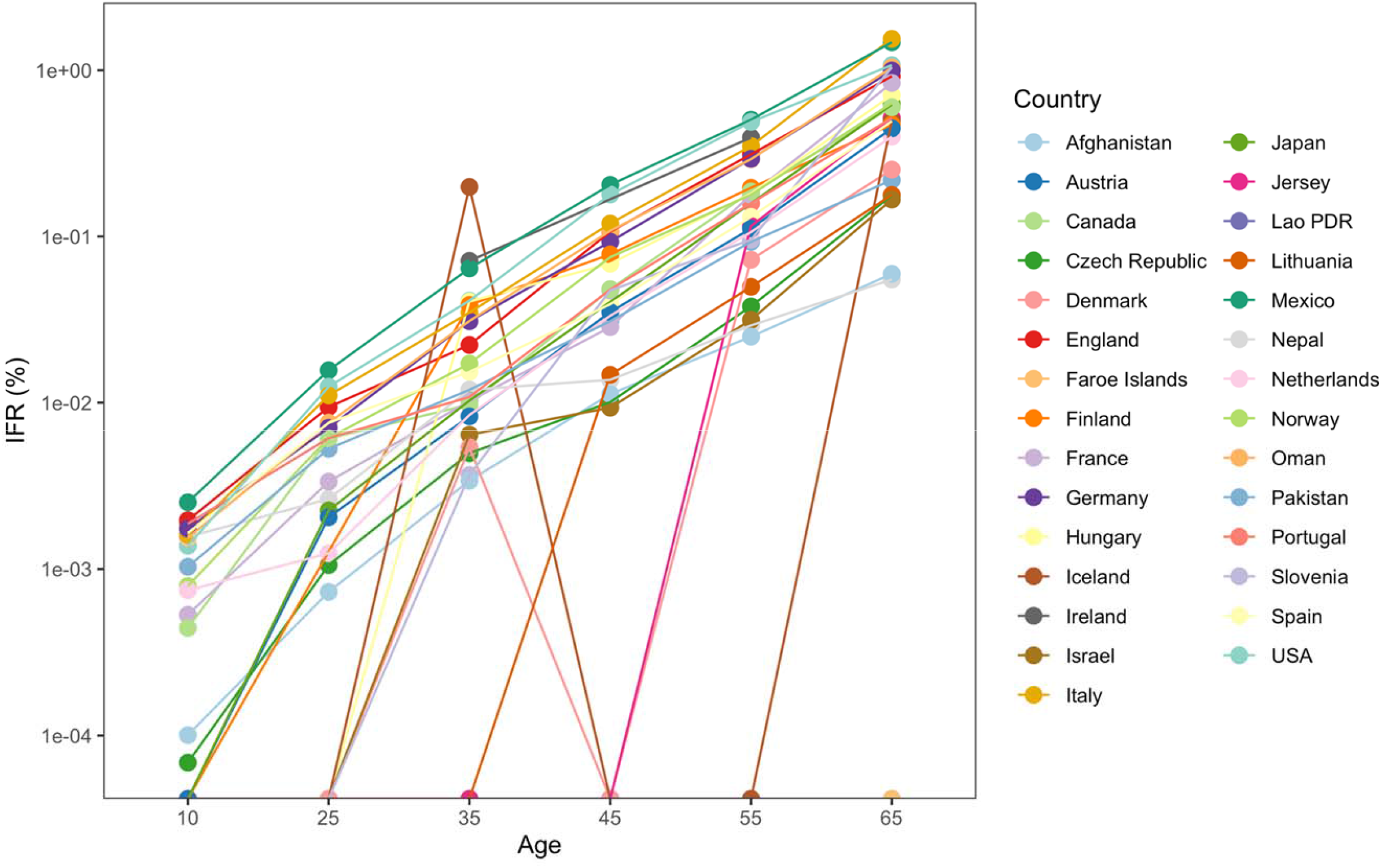
IFR in each country per each specified age bin.

### 3.5 Sensitivity analyses

Among high-income countries, the median IFR was 0.038% in the 0-59 years old age group and 0.098% in the 0-69 years old age group. Sensitivity analysis considering deaths up to 2 weeks after the midpoint of seroprevalence sampling, instead of just one week, yielded largely similar results (not shown). Sensitivity analysis excluding studies where the chosen time point of peak seroprevalence was not the latest available (observed seroprevalence has declined subsequently) yielded median IFR of 0.035% in the 0-59 years old group and 0.093% in the 0-69 years old age group. Appendix Table 2 shows results with different assumptions about seroreversion.

In the post hoc sensitivity analysis aiming to include all countries in the calculations, for countries without available age-stratified mortality data, 10-60% and 20-90% of COVID-19 deaths were assumed to have occurred among 0-59 and 0-69 year old people, respectively. Moreover, since data on age stratified deaths for Austria had been collected but the seroprevalence study report did not describe age stratified seroprevalence, we considered the overall seroprevalence (4.7%) for 0-59 and 0-69 age groups in this additional analysis. Under the minimum age-stratified mortality scenario, the median IFRs were 0.025% (IQR 0.006 - 0.043%) for the 0-59 and 0.063% (IQR 0.011 - 0.113%) for the 0-69 age group. Under the maximum scenario, the median IFRs were 0.032% (IQR 0.012 - 0.053%) for the 0-59 and 0.082% (IQR 0.034 - 0.117%) for the 0-69 age group.

### 3.5 Evaluation of heterogeneity

The pre-specified regression of IFR for the 0-59 years old age group against the proportion of people <50 years old (Figure 3A) had a slope of -0.002 (p□=L0.08), suggesting an IFR of 0.054%, 0.043%, and 0.026% when the proportion of people <50 years old in the 0-59 group was 77.5%, 82.5%, and 90%, respectively. The same analysis for the 0-69 years old age group (Figure 3B) had a slope of -0.004 (p□=□0.01), suggesting an IFR of 0.139%, 0.117%, 0.072%, and 0.027% when the proportion of people <50 years old in the 0-69 group was 65%, 70%, 80%, and 90%, respectively.

**Figure 3.**
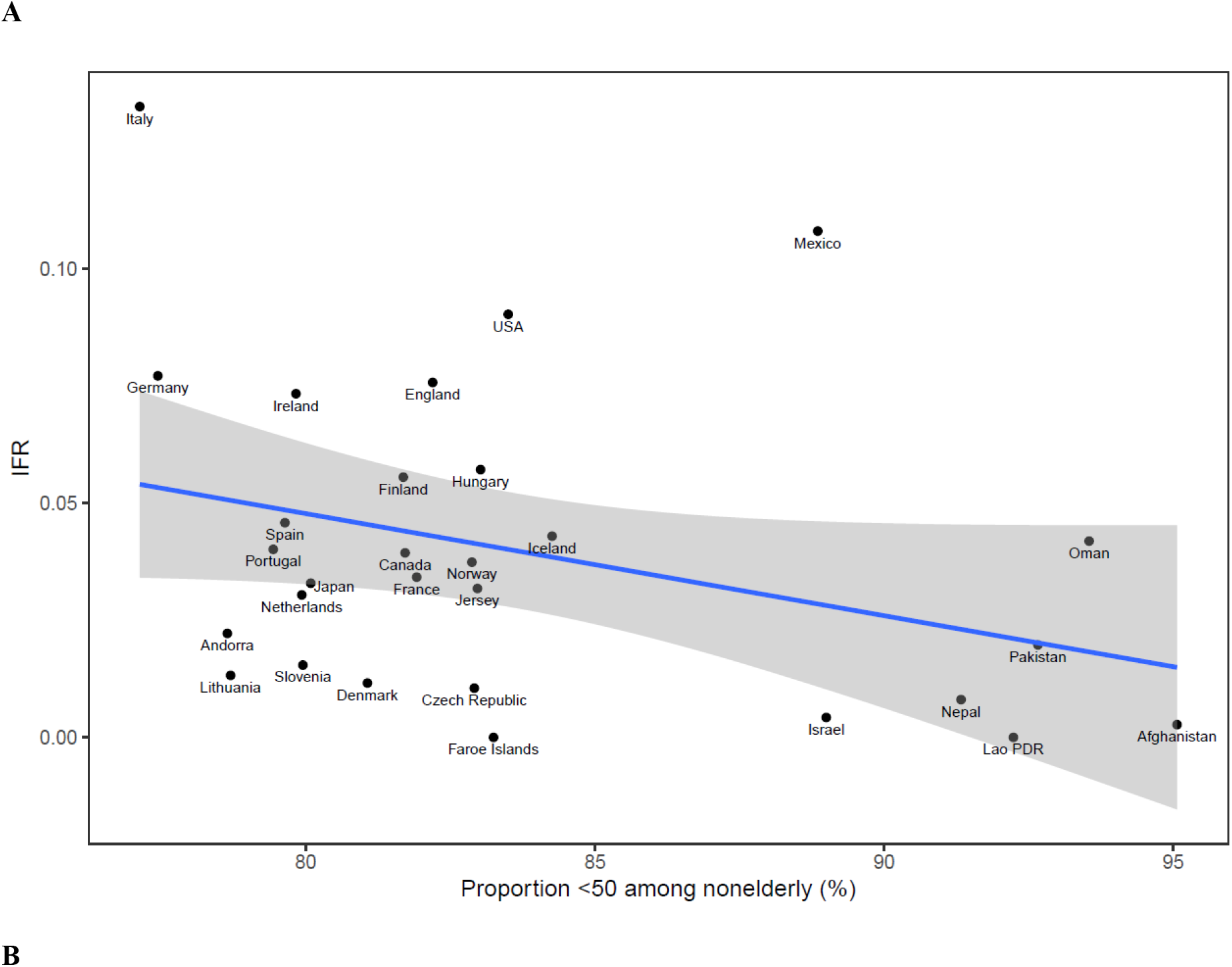
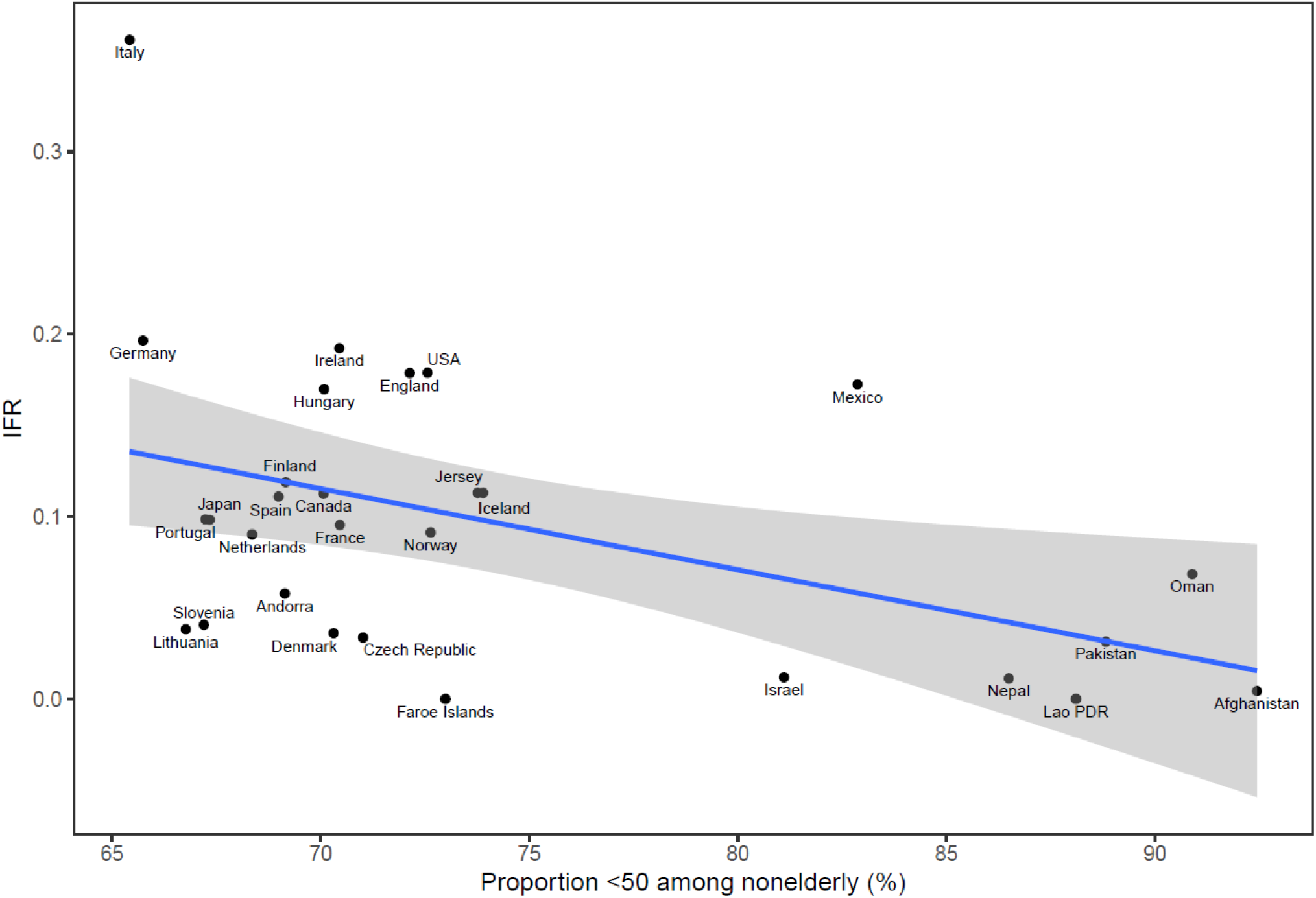
Meta-regressions of IFR as a function of the proportion of the population <50 years old among (A) among those 0-59 years old and (B) among those 0-69 years old.

The median IFR for the 0-59 years old age group was 0.038% in high-income countries versus 0.008% in other countries (p = 0.12 by Mann-Whitney U test). The median IFR for the 0-69 years old group was 0.098% in high-income countries versus 0.012% in other countries (p = 0.04 by Mann-Whitney U test). A regression of IFR for the 0-59 years old age group against the crude death rate per 1,000 people (of all ages) in each country had a slope of 0.002 (p = 0.46), while for the 0-69 age group the slope was 0.009 (p = 0.16).

## 4. DISCUSSION

The current comprehensive systematic evaluation of national seroprevalence studies suggests that the IFR of COVID-19 among non-elderly populations in the pre-vaccination era is substantially lower than previously calculated (4-8,59), especially in the younger age strata. Median IFRs show a clear age-gradient with approximately 3-4-fold increase for each decade but it starts from as low as 0.0003% among children and adolescents and it reaches 0.5% in the 60-69 years old age group. Sensitivity analyses considering all 38 countries with seroprevalence data that were identified in our systematic search showed that median IFR might be up to a third lower than the estimates produced by our main analysis, e.g. approximately 0.03% in the 0-59 years age group and 0.06-0.08% in the 0-69 years old group. Consistent with these estimates, meta-regressions suggest IFR estimates in that range for the global population where 87% of the 0-59 years old people are <50 years old and 80% of the 0-69 years old people are <50 years old.

Our IFR estimates tend to be modestly to markedly lower than several previous calculations (4-8, 59). The most comprehensive prior evaluation of COVID-19 IFR in the pre-vaccination era (59) suggested a trough IFR at the age of 7 years (0.0023%, 95% uncertainty interval 0.0015–0.0039) and increasing exponentially through 30 years (0.0573%, 0.0418– 0.0870), 60 years (1.0035%, 0.7002–1.5727) and older ages. Conversely, our median IFR estimates are roughly 10-fold lower than these previous calculations among children and young adults and 3-6-fold lower among adults 40-69 years old. If we exclude study data from age bins with 0 deaths in our calculations (a justifiable choice, since these estimates of 0% IFR are clearly underestimates), our age-stratified IFR are still approximately 2-5-fold lower than those of (59) across the entire age range. The previous IFR calculations (4-8, 59) were based on more limited national representative studies’ data and also included data from non-national samples with potentially larger bias. They also probably included mostly hard hit countries that may tend to have the highest IFR estimates. While much of the diversity in IFR across countries is explained by differences in age structure (59), additional substantial differences are possible. Another major reason for the discrepancy versus prior calculations is due to the fact that some previous calculations (e.g. ref. 59) have substantially increased their initial IFR estimates by multiplying them for a factor of under-ascertainment of COVID-19 deaths. Aligning evaluations in terms of this methodological difference would bring the estimates closer, but divergence would still be present with our estimates remaining lower. Some other estimates for pre-vaccination IFR agree more with our estimates overall, e.g. 0.107% across all ages combined (60).

The median IFR estimates should not diminish attention to the large heterogeneity that was observed across different studies and countries. Some of the observed heterogeneity may be data artefacts (e.g. if the number of deaths or seroprevalence are not accurately measured) and some may reflect genuine differences across populations and settings. Fatality risk from COVID-19 is strongly influenced by the presence and severity of comorbidities (61). While this is extremely well documented from population studies, IFR estimates stratified for comorbidity are typically not available in national seroprevalence studies. A national study of blood donors in Denmark has estimated an IFR of only 0.00336% for people < 51 years without comorbidity, and 0.281% for people aged 61-69 years old without comorbidity (62). The proportion of people with some comorbidities that are very influential for COVID-19 outcomes such as obesity is very different across different countries, even for the same age groups. For example, obesity affects 42% of the USA population (63), but the proportion of obese adults is only 2% in Vietnam, 4% in India and <10% in most African countries (64). However, also within Africa, obesity affects 0% of Ethiopian women and almost 40% of South African women (65). Another influential difference is the presence of frail individuals in long-term facilities, where IFRs may be much higher and to what extent these highly vulnerable individuals are infected. Even though the vast majority of frail individuals in long-term care are ≥70 years old, a small proportion are younger and they may account for a substantial proportion of deaths in the non-elderly strata that we examined in the current analysis, especially in some high income countries, but not in others. Other differences in management, health care, overall societal support and concomitant epidemics, e.g. drug overdose (66), may have also shaped large differences across countries.

Some limitations should be acknowledged in this work. Data artefacts in the form of measurement errors may have affected the results of some studies included in this analysis, and therefore also the data synthesis. Seroprevalence studies have many caveats (7) and uncertainty in seroprevalence estimates is larger than conveyed by typical 95% confidence intervals. Overall, however, there is no reason to suggest that over-estimation of seroprevalence is far more or far less common that under-estimation. Among the 40 studies in our evaluation, the Italian national seroprevalence study provided estimates that are very far from any other study. A notable difference that we found in this study is the requirement to isolate after a positive result to the antibody test (67,68). This might have discouraged the participation of people that expected to test positive, thus likely overestimating the IFR (68). Outliers are more suspect for bias and inaccuracies, hence, we primarily focused on the median values. For death counts, it is more likely that COVID-19 deaths were under-counted in the first waves, but both over- and under-counting may have occurred to some extent in different settings (17). Some of the studies that suggest higher estimates of IFR use large corrections for under-counting of deaths (59,69).

However, it is unclear whether such large corrections are justified. In particular, for the non-elderly age groups, deaths among young adults and children may be less likely to have been missed, as opposed to deaths of elderly individuals where causal attribution to a single cause can be more difficult and where even in high income countries under-reporting of COVID-19 may have occurred if testing was not widespread. For example, in the Netherlands, the national statistics service suggests that many COVID-19 deaths may have not been recorded in the first wave; however, these pertained largely to elderly individuals (70).

Consistent with the very low IFR estimates in non-elderly that we have obtained in this work, excess death calculations (71) show no excess deaths among children and adolescents during the pandemic in almost any country that has highly reliable death registration data. In most of these countries, moreover, excess deaths in non-elderly adults are very limited, but exceptions do occur, most notably in the USA where almost 40% of excess deaths were in populations younger than 65 years (71). This picture is very consistent with the overall very low IFR in the non-elderly, but also the large diversity in the risk profiles of populations in different countries.

Finally, the data that we analyzed pertain to the pre-vaccination period. During 2021 and 2022, the use of vaccination and the advent of new variants plus pre-existing immunity from prior infections resulted in a marked decline in the IFR. Studies in Denmark (72) and Shanghai (73) suggest that in 2022, IFRs in vaccinated, previously not infected populations were extremely low. For example, in Denmark, IFR was only 1.6 per 100,000 infections for ages 17-35 and even in ages 61-72 it was only 15.1 per 100,000 infections. In Shanghai, in 2022, IFR was 0.01% among vaccinated individuals aged 40-59 and close to 0% for younger vaccinated people, while it was practically 0% for children and adolescents regardless of vaccination. Other population studies, e.g. in Vojvodina, Serbia (74), suggest that fatality rates may be ten times lower in re-infections versus primary infections. The relative contributions of vaccination, prior infection and new variants in the IFR decline needs careful study and continued monitoring. However, it is reassuring that even in the wild strains that dominated the first year of the pandemic, the IFR in non-elderly individuals was much lower than previously thought.

## Data Availability

The protocol, data, and code used for this analysis will be made available at the Open Science Framework upon publication: https://osf.io/xvupr

## Credit author statement

A.M.P.: Conceptualization; Data curation; Formal analysis; Investigation; Writing – original draft; Writing – review & editing. C.A.: Conceptualization; Data curation; Investigation; Writing – review & editing. D.G.C.-I.: Conceptualization; Data curation; Investigation; Writing – review and editing. A.A.: Conceptualization; Data curation; Investigation; Writing – review and editing. J.P.A.I.: Conceptualization; Data curation; Formal analysis; Investigation; Writing – original draft; Writing – review & editing.

## Ethical approval

Not applicable.

