## Supplementary material for "Age-stratified infection fatality rate of COVID-19 in the non-elderly informed from pre-vaccination national seroprevalence studies"

*equal first authors

^1^Meta-Research Innovation Center at Stanford (METRICS), Stanford University, Stanford, California, USA

^2^Sezione di Igiene, Dipartimento di Scienze della Vita e Sanità Pubblica, Università Cattolica del Sacro Cuore, Rome, Italy

^3^Division of Infectious Diseases, Department of Pediatrics, Stanford University School of Medicine, Stanford, California, USA

^4^Faculty of Medicine, Université de Montréal, Montreal, Canada

^5^Departments of Medicine, of Epidemiology and Population Health, of Biomedical Data Science, and of Statistics, Stanford University, Stanford, California, USA

Correspondence to: John P. A. Ioannidis, Stanford Prevention Research Center, Medical School Office Building, Room X306, 1265 Welch Road, Stanford CA 94305, USA

**Appendix Figure 1.** Flowchart of the search and selection process.

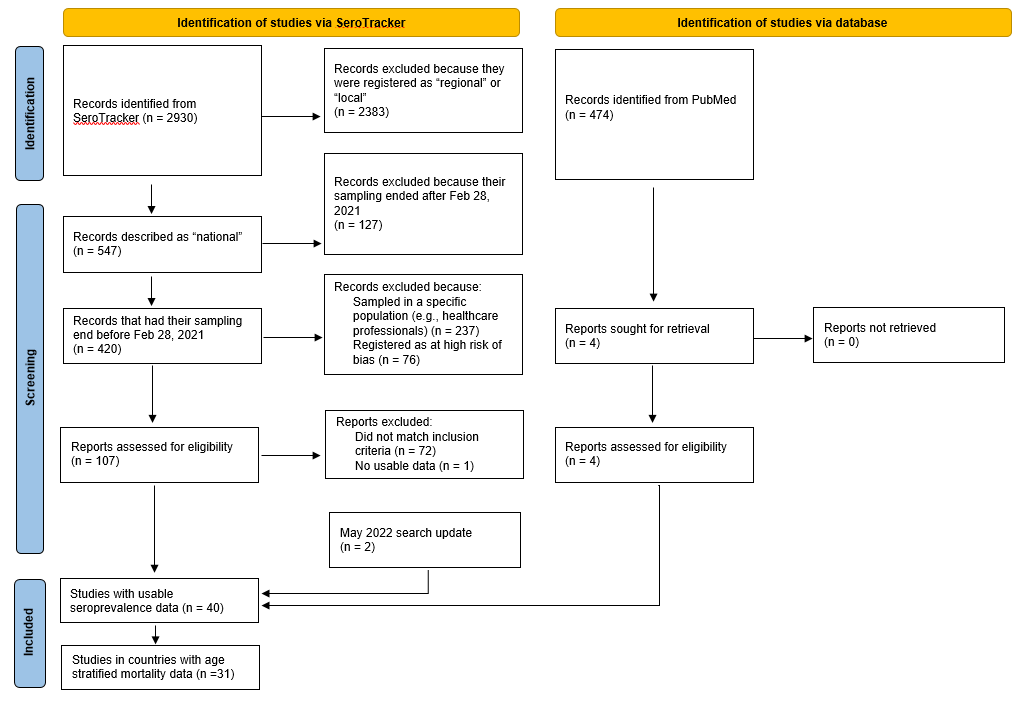

Note. In a parallel project on differential seroprevalence rates included in the same protocol as this study (preprint: https://doi.org/10.1101/2022.06.28.22277034), the flowchart is largely similar, but two more SeroTracker reports among the 107 assessed for eligibility were excluded for no usable data, since its analysis necessitated seroprevalence data for more than one age group. That was not needed here, and so 34/107 SeroTracker reports had usable data. Reasons for exclusion are given in the parallel project preprint’s appendix.

**Appendix Table 1:** Amendments/clarifications to protocol.

| Manuscript section | Amendment/Clarification | Rationale |
| --- | --- | --- |
| Eligible seroprevalence studies | For separating seroprevalence of non-elderly adults from elderly, we accepted cut-offs in the range of 54-70, preferring the one available that was closest to 60. | For elderly cut-off accepted also one study (Portugal) with cut-off at 54 years (its exclusion gives similar results). |
| Extracted information | For Mexico, we used the number of deaths according to the national surveillance system because these mortality data have been used to calculate unbiased estimates of IFR by the same team that published the Mexico seroprevalence with age-specific data about the number of deaths for Mexico. The data count the deaths in each region a month after the region-specific median date of the sampling. | Data accuracy |
| Extracted information | In Wikipedia, the article on COVID-19 in Iceland gives the specific ages of all the first victims: among the 3 deaths <70, 1 of them is <60, so we used 1 death to calculate IFR in the 0-60 age group. For the narrow age bins, we used 1 death 30-39, 2 deaths 60-69. | Data availability and accuracy |
| Extracted information | For Ireland we did not have for deaths a 10-year interval available for imputation. The publication of the seroprevalence study states that there were 190 COVID-19 deaths 12-69 years old by July 16, 2020. As per protocol, we can use these 190 deaths as the numerator for the IFR calculation in the 0-70 estimate. We found from government sources n=110 deaths for 0-64 years. We can approximate the deaths in the 0-60 assuming a similar step decrease for the 5 years from 65 to 60 as from 70 to 65. This means that the deaths in 0-70 are 190 and the deaths in 0-60 are 64. | Data availability |
| Extracted information | Deaths for <70 Andorra are imputed from the previous project, but we have no data for 0-60. We will use the median ratio of deaths 0-70/ deaths0-60 in all other countries which is about 3, and thus assume that for Andorra the number of deaths 0-60 is 2. | Data availability |
| Extracted information | Kalish (USA) explicitly excluded in their sampling for SP testing all the previously detected case. So, when we estimated the number of infected people we needed to add those cases. Since they found 4.8 undiagnosed cases for each diagnosed case, we multiplied the number of infected that we got from the seroprevalence by 1.21-fold (5.8/4.8). | Clarification/specification needed |
| Extracted information | Sullivan and colleagues (USA) calculated that the seroprevalence adjusted for waning antibodies at October 30, 2020 is 11.9%. We used this estimate to adjust age-stratified seroprevalence reported in their study to obtain seroprevalence estimates adjusted for seroreversion. Without adjustment for seroreversion, the SP is 2.48% in the >65, 5.55% for the <65, and 4.71% for the total. Therefore, what we used for Sullivan for the non-elderly seroprevalence is 11.9x5.55/4.71=14.0%. For all the narrower age bins, similarly we multiplied the seroprevalence by a factor of 14/4.71=2.97, which is a fair approximation, assuming that the effect of waning is the same proportionally across all age strata. | Clarification/specification needed |
| Extracted information | For the number of infected people in Iceland we used the estimates provided by the Authors in table S7 (Supplementary materials) | Clarification/specification needed |
| Extracted information | Worldometer has no data for England, the relevant data (number of deaths at the preferred date, number of deaths a week after the preferred date, mortality peak) have been retrieved from: https://coronavirus.data.gov.uk/details/deaths?areaType=nation&areaName=England | Data availability |
| Extracted information | Worldometer has no data for Jersey (Channel Islands), the relevant data (number of deaths at the preferred date, number of deaths a week after the preferred date, mortality peak) have been retrieved from: https://opendata.gov.je/dataset/coronavirus-covid-19-number-of-cases-in-jersey | Data availability |
| Extracted information | World Population Review lists Andorra annual mortality as 0 per 1000. We retrieved the correct annual mortality that is reported from multiple sources (World Bank, Statista, Knoema) as 3.9 per 1000 in 2019. | Data accuracy |
| Sensibility analyses | We added a sensitivity analysis where we included in the overall calculations of IFR in the nonelderly the data from countries where the proportion of COVID-19 deaths occurring among the non-elderly was not available. Specifically, we assumed that the proportion of COVID-19 deaths represented by the non-elderly was a minimum of 10% for 0-59 years and 20% for 0-69 years and a maximum of 60% for 0-59 years and 90% for 0-69 years. | We had data from 40 seroprevalence studies from 38 countries. We did not have age stratified mortality data for eight countries (namely, Maldives, Senegal, Russia, Mongolia, Lebanon, Jordan, Iran, India). For Austria, we had age-stratified mortality data, but not enough age stratified seroprevalence data (only 16-24, 24+, overall). To see how our findings were affected if we included all the data: (1) for the countries without age stratified mortality data, we considered the proportion of deaths under a minimum and maximum scenario; (2) for Austria, we assumed that the overall seroprevalence did not change across all age strata. |
| IFR estimates in the non-elderly | The presented 95% confidence intervals for seroprevalence and IFRs are estimated primarily as direct extractions from the seroprevalence studies. For studies that did not report 95% confidence intervals, we complemented with a calculation using the number of sampled and seropositive individuals. For those that provided adjusted estimates for age brackets (e.g., 0–9, 10–19, 20-29, etc.), we combined estimates for each study using a fixed effects inverse variance meta-analysis (of arcsine transformed proportions) to obtain 95% CIs. Asymmetry to point estimates may be observed for these cases, since point estimates were calculated by multiplying age bracket seroprevalence by the corresponding population count (which is preferable, since it takes into account population distribution) | Clarification/specification needed |

**Appendix Table 2.** Uncorrected and corrected for seroreversion infection fatality rate in non-elderly

| **Location (First author)** | **Study midpoint** | **Peak of wave 1** | **Time lag between primary date and peak (months)** | **IFR (%) uncorrected for seroreversion <60 [<70]** | **IFR (%) corrected for 1% relative seroreversion/month <60 [<70]** | **IFR (%) in non-elderly, corrected for 5% relative seroreversion/month <60 [<70]** | **IFR (%) in non-elderly, corrected for 10% relative seroreversion/month <60 [<70]** |
| --- | --- | --- | --- | --- | --- | --- | --- |
| Afghanistan (Saeedzai) | 15/06/20 | 20/07/20 | -0.69 | 0.003 [0.004] | 0.003 [0.004] | 0.003 [0.004] | 0.003 [0.004] |
| Andorra (Royo-Cebrecos) | 16/05/20 | 09/04/20 | 1.68 | 0.026 [0.068] | 0.024 [0.062] | 0.022 [0.058] | 0.02 [0.053] |
| Canada (Tang) | 16/07/20 | 06/05/20 | 2.79 | 0.047 [0.133] | 0.044 [0.126] | 0.039 [0.113] | 0.034 [0.097] |
| Czech Republic (Piler) | 31/12/20 | 14/04/20 | 9.03 | 0.017 [0.053] | 0.015 [0.049] | 0.011 [0.034] | 0.006 [0.021] |
| Denmark (Espenhain) | 16/12/20 | 09/04/20 | 8.71 | 0.018 [0.056] | 0.017 [0.052] | 0.012 [0.036] | 0.007 [0.023] |
| England (Ward) | 01/07/20 | 09/04/20 | 3.19 | 0.089 [0.21] | 0.086 [0.204] | 0.076 [0.179] | 0.064 [0.15] |
| Faroe Islands (Petersen) | 25/11/20 | NA* | NA* | 0 [0] | 0 [0] | 0 [0] | 0 [0] |
| Finland (Melin) | 03/09/20 | 26/04/20 | 4.73 | 0.071 [0.151] | 0.068 [0.144] | 0.056 [0.119] | 0.043 [0.092] |
| France (Warszawski) | 24/11/20 | 08/04/20 | 8.02 | 0.059 [0.164] | 0.054 [0.151] | 0.039 [0.109] | 0.025 [0.07] |
| France (Carrat) | 17/07/20 | 09/04/20 | 3.71 | 0.041 [0.112] | 0.033 [0.092] | 0.029 [0.079] | 0.023 [0.065] |
| Germany (Neuhauser) | 11/11/20 | 18/04/20 | 7.26 | 0.077 [0.196] | 0.077 [0.196] | 0.077 [0.196] | 0.077 [0.196] |
| Hungary (Merkely) | 08/05/20 | 23/04/20 | 0.95 | 0.06 [0.178] | 0.059 [0.177] | 0.057 [0.17] | 0.054 [0.161] |
| Iceland (Gudbjartsson) | 16/05/20 | 10/04/20 | 1.64 | 0.038 [0.1] | 0.046 [0.121] | 0.043 [0.113] | 0.039 [0.103] |
| Ireland (Heavey) | 04/07/20 | 25/04/20 | 2.76 | 0.09 [0.236] | 0.082 [0.215] | 0.073 [0.192] | 0.063 [0.166] |
| Israel (Reicher) | 09/07/20 | 18/08/20 | -0.85 | 0.006 [0.015] | 0.004 [0.012] | 0.004 [0.012] | 0.004 [0.012] |
| Italy (Sabbadini) | 19/06/20 | 03/04/20 | 2.99 | 0.157 [0.421] | 0.152 [0.409] | 0.135 [0.361] | 0.115 [0.307] |
| Japan (Yoshiyama) | 04/06/20 | 05/05/20 | 1.45 | 0.035 [0.106] | 0.035 [0.104] | 0.033 [0.098] | 0.03 [0.091] |
| Jersey (Government of Jersey) | 16/06/20 | 30/04/20 | 2 | 0.035 [0.125] | 0.035 [0.123] | 0.032 [0.113] | 0.029 [0.101] |
| Lao PDR (Virachith) | 03/09/20 | NA** | NA** | 0 [0] | 0 [0] | 0 [0] | 0 [0] |
| Lithuania (Smigelskas) | 25/08/20 | 18/04/20 | 4.7 | 0.018 [0.053] | 0.016 [0.046] | 0.013 [0.038] | 0.01 [0.03] |
| Mexico (Basto-Abreu) | 30/09/20 | 21/07/20 | 2.79 | 0.125 [0.199] | 0.121 [0.193] | 0.108 [0.172] | 0.093 [0.148] |
| Nepal (Government of Nepal) | 16/10/20 | 31/08/20 | 1.97 | 0.009 [0.013] | 0.009 [0.012] | 0.008 [0.011] | 0.007 [0.01] |
| Netherlands (Vos) | 14/06/20 | 07/04/20 | 2.69 | 0.035 [0.104] | 0.034 [0.101] | 0.03 [0.09] | 0.026 [0.078] |
| Norway (Eik Anda) | 20/12/20 | 08/04/20 | 8.87 | 0.059 [0.144] | 0.054 [0.131] | 0.037 [0.091] | 0.023 [0.056] |
| Oman (Al Abri) | 10/11/20 | 03/08/20 | 3.71 | 0.051 [0.083] | 0.049 [0.08] | 0.042 [0.068] | 0.034 [0.056] |
| Pakistan (Ahmad) | 30/10/20 | 21/06/20 | 4.76 | 0.025 [0.04] | 0.024 [0.038] | 0.02 [0.031] | 0.015 [0.024] |
| Portugal (Canto e Castro) | 14/09/20 | 14/04/20 | 5.49 | 0.053 [0.13] | 0.05 [0.123] | 0.04 [0.098] | 0.03 [0.073] |
| Slovenia (Poljak) | 29/10/20 | 12/04/20 | 7.03 | 0.022 [0.058] | 0.021 [0.054] | 0.015 [0.041] | 0.011 [0.028] |
| Spain (Government of Spain) | 22/11/20 | 03/04/20 | 8.11 | 0.07 [0.17] | 0.064 [0.155] | 0.046 [0.111] | 0.03 [0.071] |
| USA (Sullivan) | 30/10/20 | 21/04/20 | 6.77 | 0.077 [0.153] | 0.077 [0.153] | 0.077 [0.153] | 0.077 [0.153] |
| USA (Kalish) | 20/06/20 | 17/04/20 | 2.56 | 0.111 [0.219] | 0.108 [0.214] | 0.097 [0.192] | 0.084 [0.167] |

* No registered deaths till preferred date; ** No registered deaths in non-elderly till preferred date
